# The longitudinal role of overweight and obesity women in mammographic breast screening participation: retrospective cohort study using linked data

**DOI:** 10.1101/2024.01.08.24301020

**Authors:** K.A. McBride, S. Munasinghe, S. Sperandei, A. Page

## Abstract

**Objectives:** This study investigated the association between prospective changes in BMI and longitudinal adherence to mammographic screening among overweight or obese women residing in New South Wales, Australia

**Methods:** This was a retrospective cohort study of women participating in the Australian Longitudinal Study on Women’s Health (ALSWH), with subsequent screening participation enumerated via BreastScreen New South Wales, Australia clinical records over the period 1996-2016. The association between BMI and subsequent adherence to screening was investigated in a series of marginal structural models, incorporating a range of socio-demographic, clinical, and health behaviour confounders. Models were also stratified by proxy measures of socio-economic status (private health insurance and educational achievement).

**Results:** Participants who had overweight/obesity were more likely to be non-adherent to mammography screening, compared to normal or underweight participants (OR=1.29, [95% CI=1.07, 1.55). The association between overweight/obesity and non-adherence was slighter stronger among those who ever had private health insurance (OR=1.30, [95% CI=1.05, 1.61) compared to those who never had private health insurance (OR=1.19, [95% CI=0.83, 1.71), and among those with lower educational background (OR=1.38, [95% CI=1.08, 1.75) compared to those with higher educational background (OR=1.27, [95% CI=0.93, 1.73).

**Conclusion:** Findings show long-term impacts on screening participation with higher BMI women being less likely to participate in routinely organised breast screening. Women with a higher BMI should be a focus of efforts to improve breast screening participation, particularly given their increased risk of post-menopausal breast cancer and the association of higher BMI with more aggressive clinical presentations and histopathology of breast cancers.

**Key messages:**

1. Overweight and obesity increase risk of breast cancer, poorer prognostic features and worse outcomes.
2. Long-term impacts on screening participation are evident among higher BMI women who are less likely to participate in routinely organised breast screening.
3. This relationship is stronger among women of lower educational attainment.
4. Women with a higher BMI should be a focus of targeted efforts to improve their breast screening participation

## BACKGROUND

Breast cancer is the leading cause of cancer death among women globally, with seven women on average dying each day in Australia from the disease [1]. Breast cancer places a heavy burden on healthcare systems, with expenditure on breast cancer treatment forecast to rise in coming years due to increasing incidence, an aging population and an increase in risk factors, such as obesity [2]. Early detection and intervention for breast cancer is key in achieving better survival rates with biennial mammographic screening and earlier detection shown to be associated with a reduction in mortality from breast cancer of between 21–28% [2]. In Australia, like other developed settings, mammography screening is population-based and provided via a free organised screening program for all women aged over 40 in Australia. Women between the ages of 50-74 are routinely invited to biennial screening as part of this BreastScreen program, delivered across all states and territories, including through BreastScreen New South Wales (NSW). Women aged 40-49 can self-refer.

Despite the established benefits and availability of screening, rates of participation in Australia remain suboptimal, with approximately 55% of women (number of women screened in 2-years divided by the eligible index cohort) regularly participating in the national program in 2017-2018 [3], well below BreastScreen’s National Accreditation Standard target of ≥70% [4]. Screening participation is also lower in certain areas and among certain populations, with lower rates in rural and remote regions, among people who experience socioeconomic disadvantage, individuals from certain ethnic backgrounds, and those of Aboriginal or Torres Strait Islander status. However, these sub-populations are also at higher risk of breast cancer due to the higher prevalence of risk factors including obesity [5, 6]. Obesity is a well-established risk factor for the development of post-menopausal breast cancer due to adipose tissue acting a major reservoir for oestrogen biosynthesis following menopause [7–9]. Higher body mass index (BMI) has also been associated with more aggressive clinical presentations of breast cancer [10, 11], is an adverse prognostic factor in response to adjuvant chemotherapy [12], and is associated with higher mortality rates due to breast cancer [13, 14].

In NSW, the population focus of the current study, the BreastScreen NSW program currently has one of the lowest participation rates (53.3%, 2018-2019) of all breast-screening programs in Australia [15], and is also a population where 1 in 3 women (29.96%) in the target screening age group have obesity [16]. This suggests there are a number of higher risk women in NSW who may not be accessing timely or regular mammography screening. However, the true incidence proportion of women who choose not to screen, or do not return for their biennial screening mammogram, because of issues associated with their weight is unclear. It is also not known how weight might interact with other potential screening influencers such as cultural background or level of education.

Exploratory research conducted in NSW among women with obesity has identified possible reasons for low participation in mammographic screening [17], including poor prior screening experiences, limited desire to prioritise personal health needs and low knowledge around the heightened need to screen. Suboptimal mammographic screening participation has also been reported among women with obesity in other settings, largely established through cross sectional studies based on self-reported screening behaviour [18–20]. Only one study in Australia, based on self-reported screening data, found obese women were 8% less likely to screen than ‘healthy’ weight women [21] and may be over-estimating of the proportion of women actually screening [21]. Associations like these are useful in informing further research directions but are unable to demonstrate a clear temporal association between BMI and adherence to screening. Research is needed to examine how changes in BMI and other co-variates determine subsequent mammographic screening adherence, using objectively collected screening data and accounting for longitudinal changes in BMI. Accordingly, this study investigates the association between prospective changes in BMI and longitudinal adherence to mammographic screening among overweight or obese women residing in NSW, Australia.

## METHODS

### Study design

This was a retrospective cohort study design based on a cohort of women participating in the Australian Longitudinal Study on Women’s Health (ALSWH), with subsequent screening participation enumerated via the BreastScreen NSW clinical records. Data were linked using individual-level probabilistic record linkage by the Centre for Health Record Linkage (CheReL). Ethics approval was obtained from the NSW Population and Health Service Research Ethics Committee (2019/ETH01566).

### Data sources

#### The Australian Longitudinal Study on Women’s Health data

The ALSWH is a longitudinal population-based survey examining the health of over 58,000 Australian women in 3 cohorts who were aged 18-23, 45-50 and 70-75 when surveys began in 1996. ALSWH assesses physical and mental health, as well as psychosocial aspects of health (including socio-demographic and lifestyle factors) and use of health services among women who are broadly representative of the Australian population. Women in ALSWH were randomly selected from the Medicare Australia database, with deliberate oversampling from rural and remote areas. More than 40,000 women responded to an initial invitation, with each age cohort completing a survey every three years. The current study is based on women from the 1946-51 cohort (women now aged >= 70 years) who reside in NSW. This cohort (aged 45-50 in 1996) were resurveyed in 1998, 2001, 2004, 2007, 2010, 2013 and 2016 to provide eight waves of longitudinal data.

#### BreastScreen NSW mammography clinical records

BreastScreen NSW is part of a national mammography screening program jointly funded by the Commonwealth, State, and Territory governments. BreastScreen NSW is the central data holder for the results of all mammograms that have taken place in NSW as part of this program, and collects information on breast screening services within the program for women aged 40 years and over. For the current study, mammography screening data was obtained for clients over the period 1996-2016.

#### Data linkage

ALSWH participants were individually linked to the BreastScreen NSW clinical records using probabilistic record linkage after (i) application approval from NSW Centre for Health Record Linkage (CHeReL), (ii) application approval from the ALSWH (iii) application approval from BreastScreen NSW and (iv) institutional ethics approval from NSW Population and Health Service Research Ethics Committee (2019/ETH01566). The CHeReL is the third-party organisation in NSW that liaises with Data Custodians and conducts data linkage on behalf of researchers for the state of NSW via a Master Linkage Key (MLK) comprising identifying information including name, address, date of birth and gender.

### Study variables

The outcome variable of the study was non-adherence to routine breast screening as part of the BreastScreen program, defined as a dichotomous variable (‘yes’, ‘no’). Adherence was classified as ‘yes’ if women received a screening mammogram at BreastScreen NSW within three years of their last screening mammogram. If women did not attend for a subsequent BreastScreen mammogram within three years following their previous screening mammogram, adherence was classified as ‘no’. Women who had their last screening mammogram before December 2013 and who were still alive in December 2016 but had not returned for a subsequent screening mammogram, were also classified as ‘no’. Additionally, where the gap between two screening mammogram visits was >6 years, these cases were classified as ‘no’ for the 3-year period following the last screening mammogram and were also classified as ‘no’ for the 3-year period following the putative intervening screening mammogram that would have occurred between the two recorded screening mammogram visits.

The main exposure variable was BMI based on self-reported height and weight, classified as ‘healthy or underweight’ (BMI score <25) or ‘overweight or obese’ (BMI score >=25). Additionally, a series of time-dependent and -independent confounding variables were included to adjust for the association between BMI classification and adherence to routine breast screening (Figure 1). Potential confounders are listed in Table 1.

**Figure 1 somewhere here**

**Table 1.** Confounders.

| **Time varying confounders** | **Time invariant confounders** |
| --- | --- |
| Accessibility/Remoteness Index of Australia (ARIA) group ('major cities', 'inner regional', 'outer regional', 'remote/very remote'), marital status ('married', 'divorced', 'de-facto', 'widowed', 'separated', 'never married'), labour force status ('employed', 'unemployed', 'not in labour force'), any chronic disease ('yes', 'no'), any cancer ('yes', 'no'), any psychiatric disorder ('yes', 'no'), ease of having a mammogram ('excellent', 'very good', 'good', 'fair', 'poor', 'don't know'), smoking status ('never-smoker', 'ex-smoker', 'smoker'), ever had abnormal PAP test result ('yes', 'no', 'don't know'), ever had abnormal mammogram ('yes', 'no', 'don't know'), breast examined by a doctor or nurse ('yes', 'no'), and current hormone replacement therapy ('yes', 'no'). | Age ('45-54', '55-64', '65-74'), country of birth ('Australia born', 'Other English-speaking countries', 'Europe', 'Asia', 'Other'), educational status ('no formal education', 'school certificate', 'high school certificate', 'trade/apprentice', 'certificate/diploma', 'university degree', 'higher degree'). |

### Statistical analyses

Descriptive analyses were presented counts and percentages or mean and standard deviation, where appropriate. Since the exposure variable (BMI category) changes over time, conventional conditional statistical models may result in biased estimates of the association between the exposure and outcome, due to the over-adjustment bias or endogenous selection bias [22]. Accordingly, this study used marginal structural models (MSM) with inverse probability of treatment weights (IPTW) to provide an unbiased estimates of the controlled direct effect of BMI classification on adherence to breast cancer screening. [22, 23]

The creation of IPTW was based on two main steps. First, a propensity score model was used to derive individual probabilities of those who are exposed (‘overweight or obese’) or not exposed (‘normal or underweight’). Secondly, the weights were then calculated based on the inverse value of the individual probabilities from the first step [22, 24]. The final weight for each participant was then calculated by multiplying the individual weights calculated at each time point of the study [22]. IPTW generates a pseudo-population that is two times larger than the original population, with the distribution of confounders are equally distributed between exposed and non-exposed groups [22, 24].

When conducting an MSM analysis, there are two main types of weights (related to statistical weighting and not to BMI) including unstabilised weights and stabilised weights. In short, unstabilised weights are the inverse value of the individual probabilities derived conditional to the time-dependent exposures and previous treatment histories, whereas stabilised weights include a numerator derived based on the baseline values of the time-dependent confounders and previous exposure histories. The present study used stabilised weights to fit MSM models, adjusted and unadjusted for baseline exposure values, to examine the association between of BMI classification and adherence to breast cancer screening when specifying the model [25].

Univariable and multivariable multilevel mixed effect logistic regression (random intercept) models, were also conducted to estimate the association between BMI classification and adherence to breast cancer screening, adjusting for time-varying confounders. Stratified analyses were conducted to investigate potential effect measure modification by private health insurance status and educational achievement as proxy indicators of socio-economic position. Stratified analyses were conducted for (i) those reporting ‘ever had private health insurance’ vs ‘those never had private health insurance’, and (ii) those with ‘lower educational achievement’ (no formal education, School Certificate, High School Certificate, and trade/apprentice) vs ‘higher educational achievement’ (attained certificate/diploma, university degree or higher degree).

The present study imputed missing variable values for survey questions from the previous or subsequent survey if there were non-missing values. Additionally, participants’ visits to BreastScreen were excluded if they did not have a survey wave completed three years before or after the scheduled breast screen. All the statistical analyses were conducted using Stata version 18 [26].

## RESULTS

At baseline, most participants included in the study (N=2,822) were married (75.8%) and born in Australia (80.6%) (Table 2). Most participants had a BMI of ≤25 kg/m^2^ (43.5%), followed by a BMI of =25 kg/m^2^ to ≤30 kg/m^2^ (32.8%), then a BMI of >=30 kg/m^2^ (22.8%). Nearly 40% of participants had attained a School Certificate qualification, and one-third a Certificate Diploma, university degree or higher degree. Nearly 46% lived in inner regional areas followed by major cities (36.5%), with the majority employed (72.6%) at the baseline. More than half (57%) had experienced any chronic disease with 14.8% reporting a personal history of cancer. Nearly 65% reported their ease of having a mammogram as either excellent or very good (Table 2).

**Table 2.**
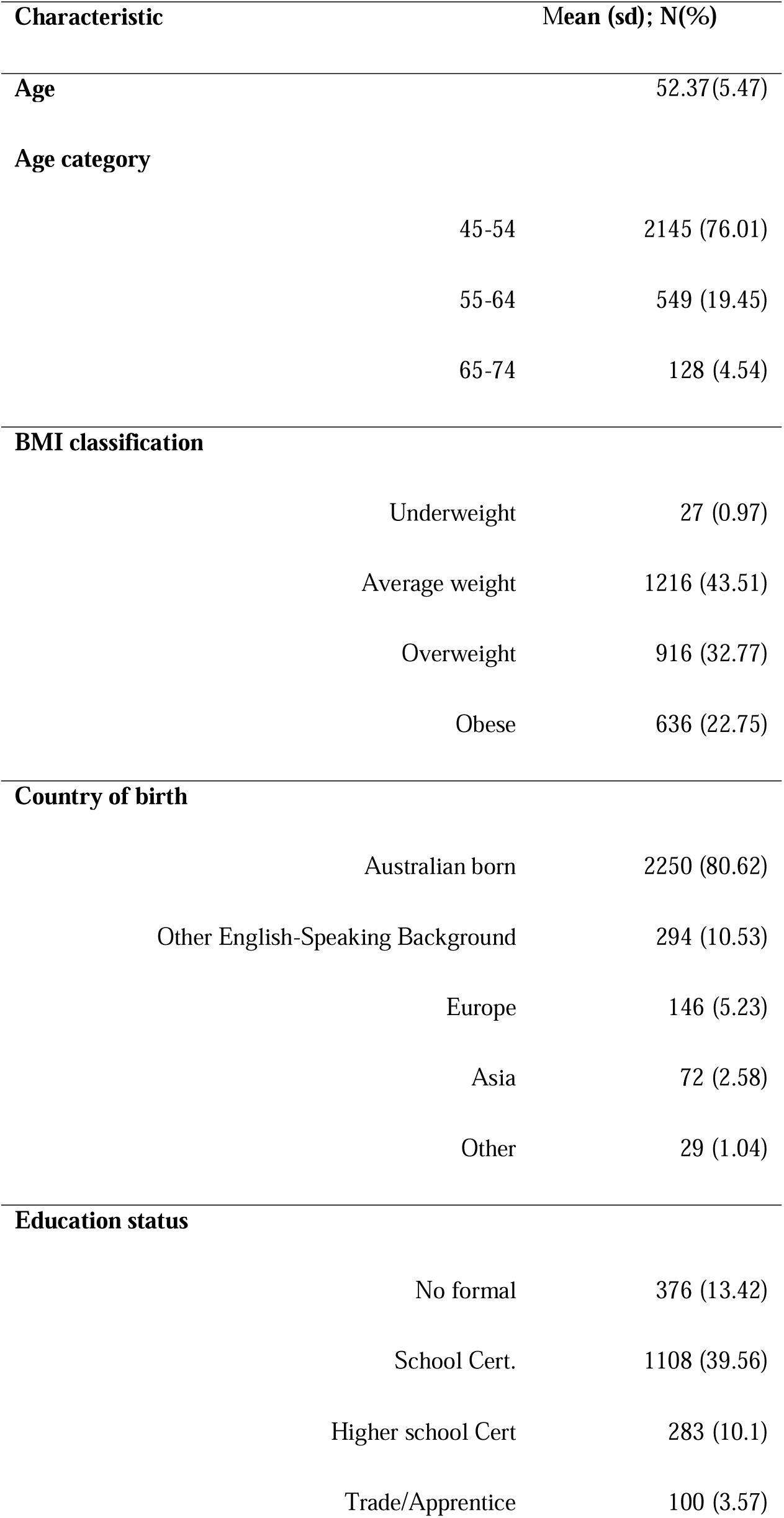
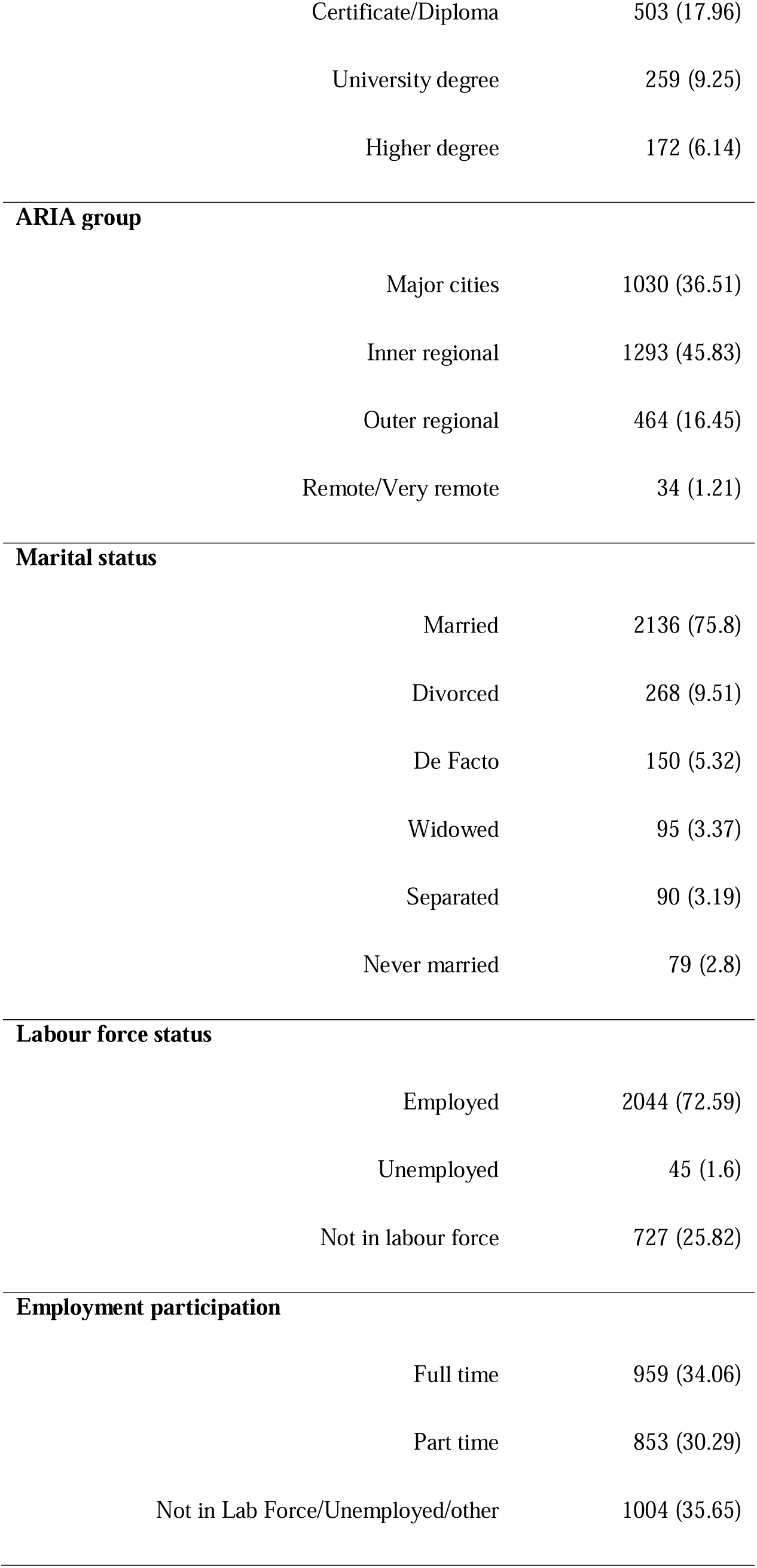
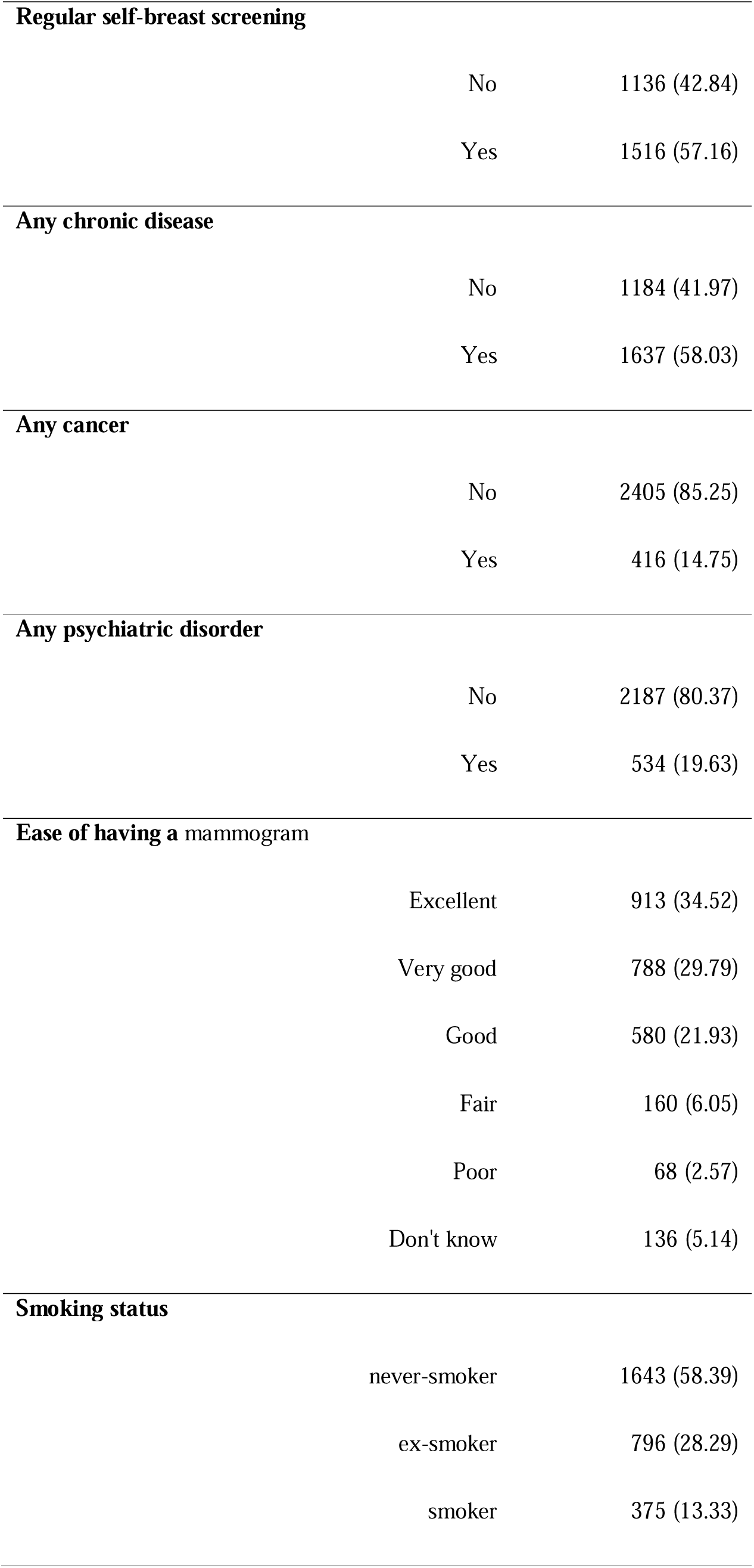
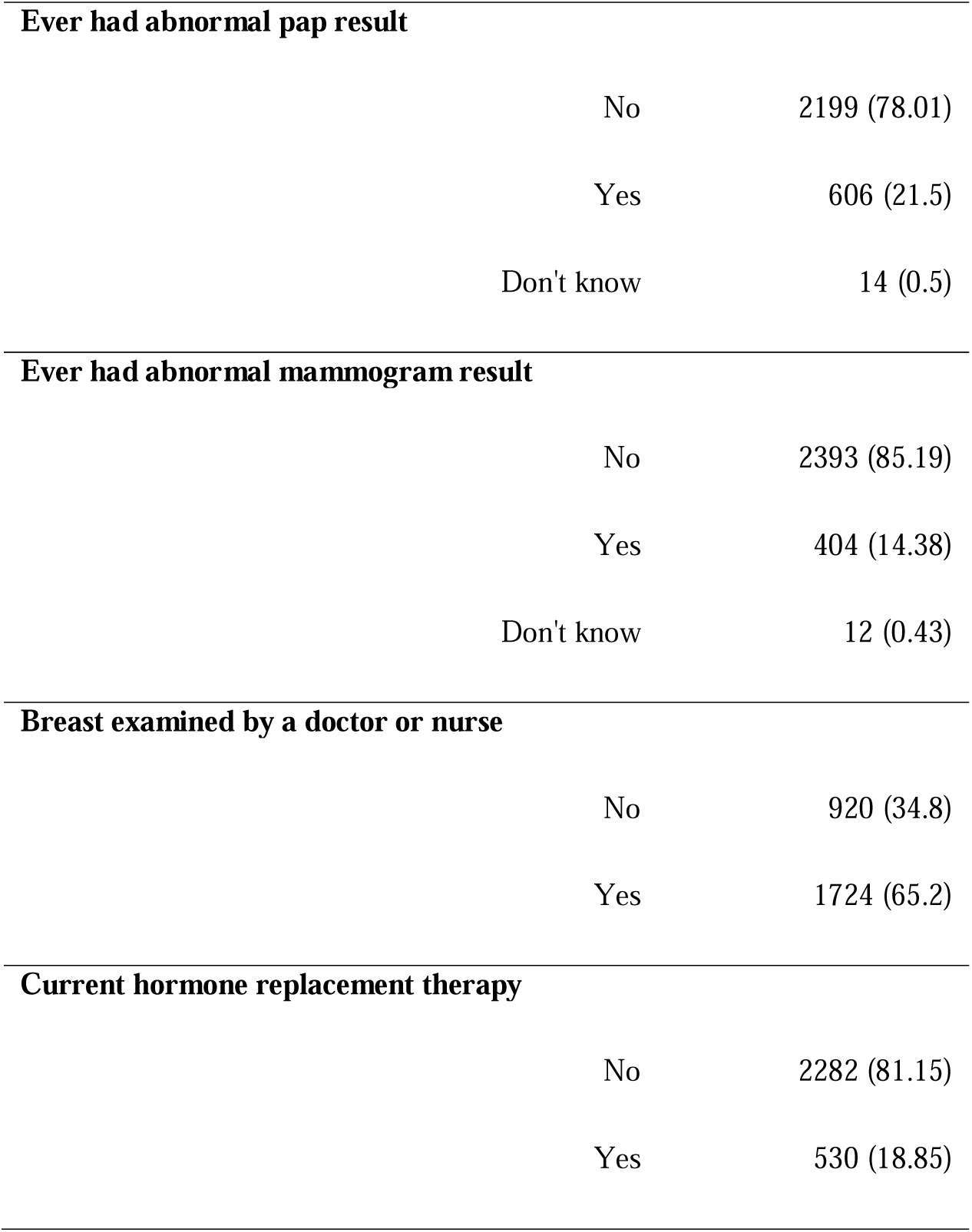
Baseline characteristics of those attended breast cancer screening (N=2822)

| Characteristic | Mean (sd); N(%) |
| --- | --- |
| **Age** | 52.37(5.47) |
| **Age category** |  |
| 45-54 | 2145 (76.01) |
| 55-64 | 549 (19.45) |
| 65-74 | 128 (4.54) |
| **BMI classification** |  |
| Underweight | 27 (0.97) |
| Average weight | 1216 (43.51) |
| Overweight | 916 (32.77) |
| Obese | 636 (22.75) |
| **Country of birth** |  |
| Australian born | 2250 (80.62) |
| Other English-Speaking Background | 294 (10.53) |
| Europe | 146 (5.23) |
| Asia | 72 (2.58) |
| Other | 29 (1.04) |
| **Education status** |  |
| No formal | 376 (13.42) |
| School Cert. | 1108 (39.56) |
| Higher school Cert | 283 (10.1) |
| Trade/Apprentice | 100 (3.57) |
| Certificate/Diploma | 503 (17.96) |
| University degree | 259 (9.25) |
| Higher degree | 172 (6.14) |
| **ARIA group** |  |
| Major cities | 1030 (36.51) |
| Inner regional | 1293 (45.83) |
| Outer regional | 464 (16.45) |
| Remote/Very remote | 34 (1.21) |
| **Marital status** |  |
| Married | 2136 (75.8) |
| Divorced | 268 (9.51) |
| De Facto | 150 (5.32) |
| Widowed | 95 (3.37) |
| Separated | 90 (3.19) |
| Never married | 79 (2.8) |
| **Labour force status** |  |
| Employed | 2044 (72.59) |
| Unemployed | 45 (1.6) |
| Not in labour force | 727 (25.82) |
| **Employment participation** |  |
| Full time | 959 (34.06) |
| Part time | 853 (30.29) |
| Not in Lab Force/Unemployed/other | 1004 (35.65) |

|  |  |  |
| --- | --- | --- |
| **Regular self-breast screening** |  |  |
|  | No | 1136 (42.84) |
|  | Yes | 1516 (57.16) |
| **Any chronic disease** |  |  |
|  | No | 1184 (41.97) |
|  | Yes | 1637 (58.03) |
| **Any cancer** |  |  |
|  | No | 2405 (85.25) |
|  | Yes | 416 (14.75) |
| **Any psychiatric disorder** |  |  |
|  | No | 2187 (80.37) |
|  | Yes | 534 (19.63) |
| **Ease of having a mammogram** |  |  |
|  | Excellent | 913 (34.52) |
|  | Very good | 788 (29.79) |
|  | Good | 580 (21.93) |
|  | Fair | 160 (6.05) |
|  | Poor | 68 (2.57) |
|  | Don't know | 136 (5.14) |
| **Smoking status** |  |  |
|  | never-smoker | 1643 (58.39) |
|  | ex-smoker | 796 (28.29) |
|  | smoker | 375 (13.33) |
| **Ever had abnormal pap result** |  |  |
|  | No | 2199 (78.01) |
|  | Yes | 606 (21.5) |
|  | Don't know | 14 (0.5) |
| **Ever had abnormal mammogram result** |  |  |
|  | No | 2393 (85.19) |
|  | Yes | 404 (14.38) |
|  | Don't know | 12 (0.43) |
| **Breast examined by a doctor or nurse** |  |  |
|  | No | 920 (34.8) |
|  | Yes | 1724 (65.2) |
| **Current hormone replacement therapy** |  |  |
|  | No | 2282 (81.15) |
|  | Yes | 530 (18.85) |

Participants who had overweight/obesity were more likely to be non-adherent to mammography screening, compared to normal or underweight participants (OR=1.29, 95% CI=1.07, 1.55) according to MSM models adjusted for baseline confounders (Table 3). Stratified analyses indicated the association between overweight/obesity and non-adherence was slighter higher among those who ever had private health insurance (OR=1.30, 95% CI=1.05, 1.61) compared to those who never had private health insurance (OR=1.19, 95% CI=0.83, 1.71) (Table 3). Similarly, stratified analyses also indicated the association between overweight/obesity and non-adherence was slightly higher among those with lower educational background (OR=1.38, 95% CI=1.08, 1.75) compared to those with higher educational background (OR=1.27, 95% CI=0.93, 1.73) (Table 3). Similar associations were evident between overweight or obese and breast screening adherence according to the multilevel mixed effect models (Supplementary Tables 1 & 2). Additionally, those more likely not to adhere to routine breast cancer screening were older (>=65 years) (OR=4.81, 95% CI=3.87, 5.98), have attained a higher degree (OR=1.74, 95% CI=1.08, 2.81), were divorced (OR=1.62, [95% CI=1.23, 2.15), had ever had a cancer (OR=1.29, 95% CI=1.09, 1.54), were current smokers (OR=1.54, 95% CI=1.15, 2.08), had ever had an abnormal mammogram test result (OR=1.71, 95% CI=1.41, 2.08), and had previously had a breast examination by a doctor or nurse (OR=1.26, 95% CI=1.08, 1.47). Individuals currently using hormone replacement therapy were more likely to adhere routine breast cancer screening (OR=0.7, 95% CI=0.56, 0.87) (Supplementary Table 1). Further, compared to those who responded as ‘excellent’ to ease of having a mammogram, those who responded ‘good’ (OR=1.53, [95% CI=1.26, 1.85), ‘fair’ (OR=3.41, [95% CI=2.51, 4.63), poor (OR=3.07, [95% CI=1.86, 5.07) and ‘don’t know’ (OR=3.63, [95% CI=2.42, 5.45) were more likely to be non-adherent to routine breast cancer screening (Supplementary Table 1). Similar associations between the above exposures and non-adherence to routine breast cancer screening were observed in multilevel models stratified by education status and private health insurance (Supplementary Table 2).

**Table 3.**
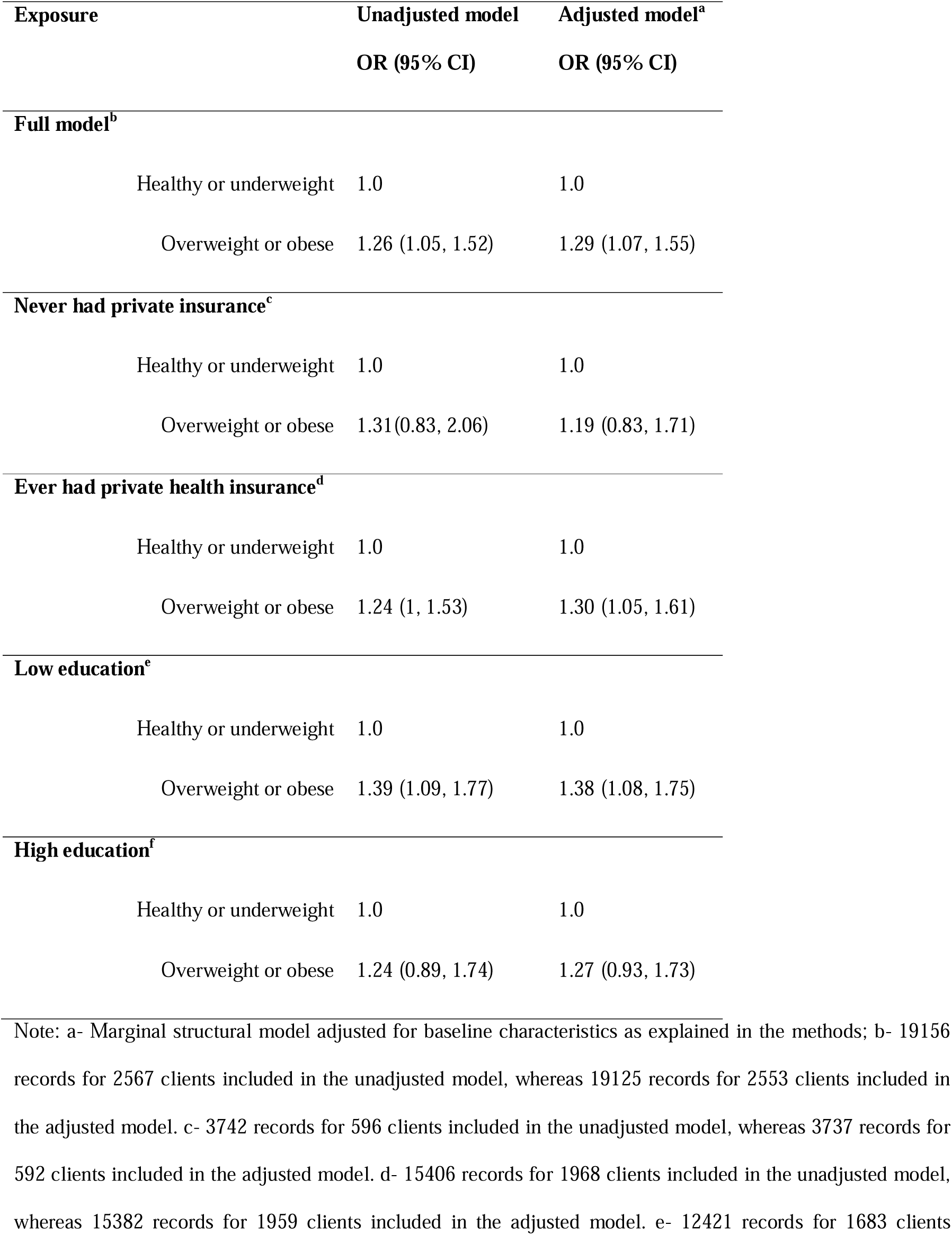
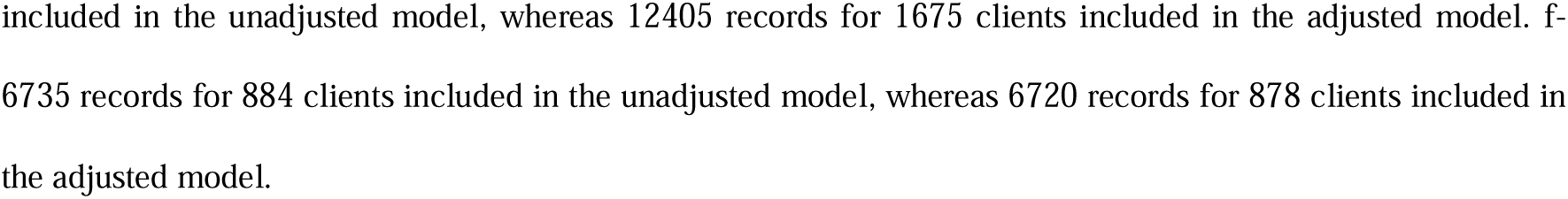
The association between body mass index category and non-adherence to mammography screening, among New South Wales women aged 50-74 years, period 1996 to period 2016 (N= 2822)

| Exposure | Unadjusted model | Adjusted model<sup>a</sup> |
| --- | --- | --- |
|  | OR (95% CI) | OR (95% CI) |
| **Full model<sup>b</sup>** |  |  |
| Healthy or underweight | 1.0 | 1.0 |
| Overweight or obese | 1.26 (1.05, 1.52) | 1.29 (1.07, 1.55) |
| **Never had private insurance<sup>c</sup>** |  |  |
| Healthy or underweight | 1.0 | 1.0 |
| Overweight or obese | 1.31(0.83, 2.06) | 1.19 (0.83, 1.71) |
| **Ever had private health insurance<sup>d</sup>** |  |  |
| Healthy or underweight | 1.0 | 1.0 |
| Overweight or obese | 1.24 (1, 1.53) | 1.30 (1.05, 1.61) |
| **Low education<sup>e</sup>** |  |  |
| Healthy or underweight | 1.0 | 1.0 |
| Overweight or obese | 1.39 (1.09, 1.77) | 1.38 (1.08, 1.75) |
| **High education<sup>f</sup>** |  |  |
| Healthy or underweight | 1.0 | 1.0 |
| Overweight or obese | 1.24 (0.89, 1.74) | 1.27 (0.93, 1.73) |

Note: a- Marginal structural model adjusted for baseline characteristics as explained in the methods; b- 19156 records for 2567 clients included in the unadjusted model, whereas 19125 records for 2553 clients included in the adjusted model. c- 3742 records for 596 clients included in the unadjusted model, whereas 3737 records for 592 clients included in the adjusted model. d- 15406 records for 1968 clients included in the unadjusted model, whereas 15382 records for 1959 clients included in the adjusted model. e- 12421 records for 1683 clients
included in the unadjusted model, whereas 12405 records for 1675 clients included in the adjusted model. f- 6735 records for 884 clients included in the unadjusted model, whereas 6720 records for 878 clients included in the adjusted model.

## DISCUSSION

This study investigated the association between BMI and subsequent participation in mammographic breast screening among a population-based cohort of NSW, Australia women. This is the first study to clearly delineate the temporal relationship between changes in BMI and longitudinal adherence to mammographic screening among women.

Our findings support the hypothesis that higher BMI has a long-term effect on participation in routinely organised breast screening, independent of individual risk factors. Women with a higher BMI should therefore be a focus of efforts to improve breast screening participation. This is particularly important given that women with a higher BMI have increased risk of post-menopausal breast cancer, more aggressive clinical presentations and histopathology of breast cancers, poorer treatment responses and metastases rates [10, 11, 27, 28]. Women with obesity have also been shown to have a one-third increased risk of breast cancer mortality and 41% increased mortality risk overall when compared to women diagnosed with breast cancer and who have a normal weight [29]. The need for focussed efforts is also highly warranted given knowledge relating to postmenopausal obesity as a risk factor for breast cancer appears to be limited [17, 30], highlighting the urgent need to educate women on their differing risk profiles.

Our findings also showed higher non-adherence to screening among women who ever had private health insurance compared to those who never had private health insurance. This could reflect misclassification bias, however, as the apparent higher non-adherence may in fact be due to women with private health insurance having mammograms outside of the BreastScreen NSW program via Medicare (the Australian Government-funded universal health insurance scheme). This type of *de facto* ‘screening’ requires general practitioner referral and requires co-payment by the patient as mammograms are partially refundable if clinically indicated, unlike the fully funded mammograms conducted via BreastScreen. *De facto* screening through alternate providers leading to late or lapsed attendance at BreastScreen has previously been associated with higher health insurance coverage, and higher education, and thought to be reflective of private screening outside publicly provided services [31–33]. Women with private health insurance (which is also a proxy for higher income) are likely to have more familiarity with the private sector and may perceive privately provided screening to be better quality.

The association between BMI and non-participation in screening was also observed to be higher among women with lower (compared to higher) educational attainment. This finding is consistent with literature examining adherence to breast screening among general populations which has found women with higher education are more likely to take part in regular breast screening when compared to women with lower education [34–37]. These findings suggest that while broad approaches aimed at all women with higher BMI may be useful, targeted, and tailored approaches to women who have a higher BMI and who also have lower education levels is warranted. This is particularly important in the context of the general lower awareness of risk breast cancer due to excess weight, given lower health literacy is likely to be a contributing factor to this issue [17, 30]. This type of approach is particularly relevant given the possibility that risk stratified screening is being trialled and considered in several settings globally underscoring the need for a well thought out approach to education [30, 38].

There are several methodological limitations when interpreting the findings from the current study. First, the baseline cohort were selected to be broadly representative of the general Australian population of women, however there was some loss to follow up in successive survey waves (from n = 13714 in 1996 to n = 7956 in 2016). At baseline, women had on average a higher BMI (54% with overweight/obesity) than similarly aged women in representative data from that time period (42% with overweight/obesity) [39]. The weight of women in the eight (final) survey wave was higher than baseline, with 63% of the remaining cohort having overweight or obesity. This proportion was lower than similarly aged women from the general Australian population (73.3%) on representative data from approximately the same time period [40], potentially under-estimating the true association between obesity and adherence to routine breast screening in the general population.

Additionally, BMI was based on self-reported estimates of height and weight, which may be a source of measurement bias in the exposure. Participants often under-estimate their weight and over-estimate their height [41].There may also be under-enumeration in the outcome measure of adherence to screening. As noted above, it is likely that a proportion of women (predominantly from higher SES groups) may have engaged in *de facto* screening through Medicare. It is not possible to determine the extent of this, however, previous estimates indicate that 36% of women who have never attended for a mammogram at BreastScreen reported a referral was required for breast screening, even though no referral is required for BreastScreen [31]. Significantly more women in this study who never attended BreastScreen screening stated they needed a doctor’s referral for screening when compared to attenders (OR 3.19, 95% CI 2.06-4.95, p<0.001). This suggests a number of women may be accessing Medicare partially funded private mammograms rather than BreastScreen mammograms.

Given the *de facto* screening may be more common among higher SES women, who are also likely to be of lower weight on average based on population estimates [40], then relative differences in adherence to screening between overweight/obese and normal weight women may be over-estimated. However, there is likely to be limited under-enumeration of subsequent BreastScreen mammograms given that all participants were able to be linked to the NSW state-wide BreastScreen clinical records up to the most recent period.

There are also some important strengths in this study. This is a large population-based cohort of women who have been followed longitudinally for 20 years allowing for the assessment of changes to BMI over time. The study also had objective enumeration of the outcome (adherence to organised breast screening) via individual-level record linkage. The analysis was also able to clearly delineate the temporal relationship between BMI and subsequent screening mammogram and incorporated a range of fixed and time-varying confounders in estimating controlled direct effects between the exposure and outcome. No previous study has investigated prospective associations between changes in BMI among women and subsequent screening behaviour.

## CONCLUSION

This study investigated the association between prospective changes in BMI and longitudinal adherence to mammographic screening among overweight or obese women residing in NSW. Findings show long-term impacts on screening participation with higher BMI women being less likely to participate in routinely organized breast screening. Women with a higher BMI should be a focus of efforts to improve breast screening participation, particularly given their increased risk of post-menopausal breast cancer and the association of higher BMI with more aggressive clinical presentations and histopathology of breast cancers.

## Data Availability

All data produced in the present work are contained in the manuscript

## ACKNOWLEDGEMENTS

The authors would like to acknowledge the Australian Longitudinal Study on Women’s Health (ALSWH) by the University of Queensland and the University of Newcastle for the survey data which was used as one of the datasets in this project. We are grateful to the Australian Government Department of Health and Aged Care for funding ALSWH and to the women who provided the survey data. We would also like to acknowledge BreastScreen NSW for provision of the other dataset used in this study, as well as the Centre for Health Record Linkage (CHeReL), NSW Ministry of Health and ACT Health, for their assistance in linking the datasets. A/Professor McBride would also like to acknowledge Western Sydney University for their support of this work through a Women’s Fellowship as well as the Women’s Health Research, Translation and Impact Network (WHRTN) for a seed grant which also supported this work.

---

**Supplementary Table 1.**
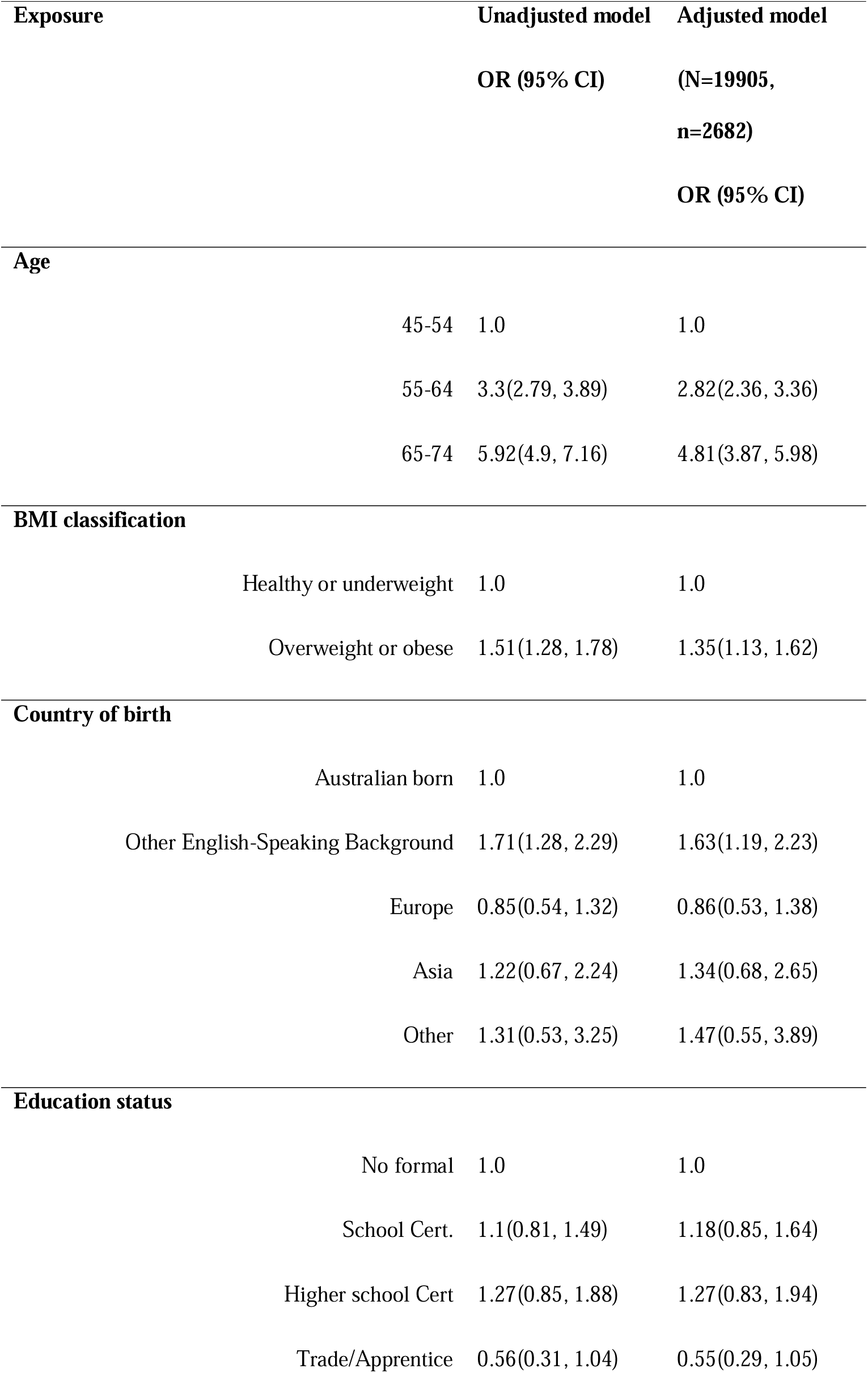
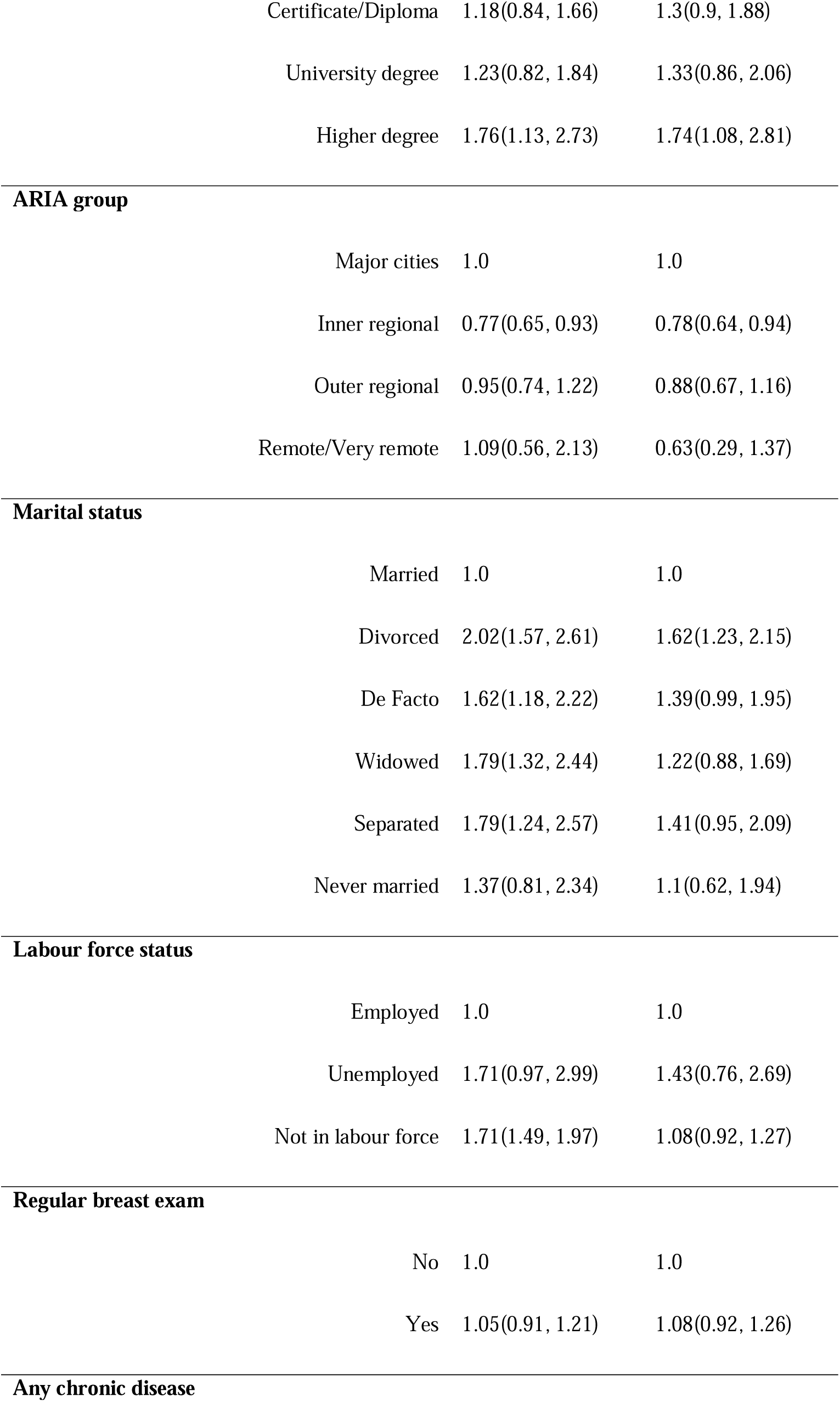
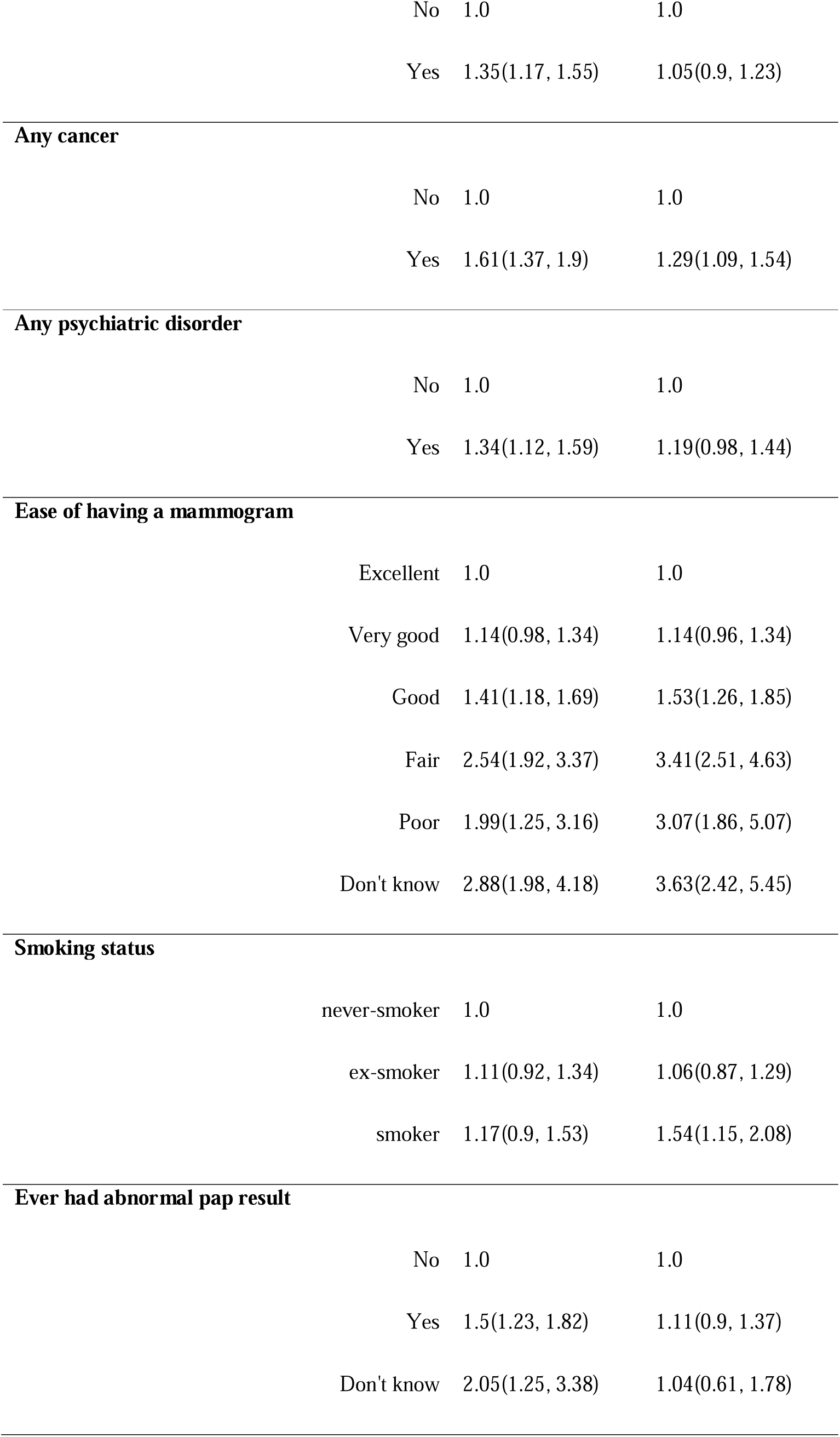
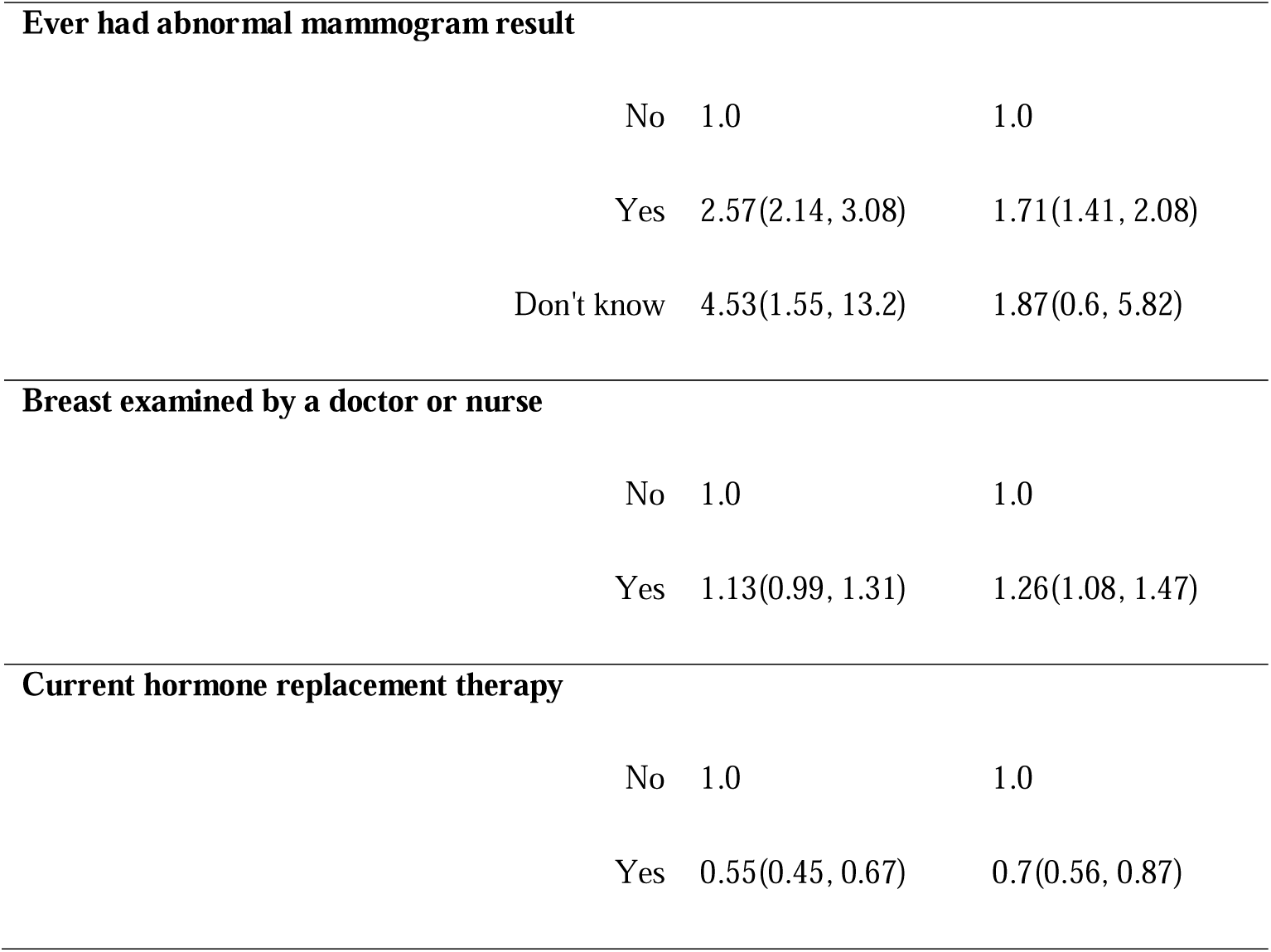
Effect of body mass index category on breast screen attendance (multilevel logistic regression approach)

| Exposure | Unadjusted model | Adjusted model |
| --- | --- | --- |
|  | OR (95% CI) | (N=19905,<br>n=2682)<br>OR (95% CI) |
| **Age** |  |  |
| 45-54 | 1.0 | 1.0 |
| 55-64 | 3.3(2.79, 3.89) | 2.82(2.36, 3.36) |
| 65-74 | 5.92(4.9, 7.16) | 4.81(3.87, 5.98) |
| **BMI classification** |  |  |
| Healthy or underweight | 1.0 | 1.0 |
| Overweight or obese | 1.51(1.28, 1.78) | 1.35(1.13, 1.62) |
| **Country of birth** |  |  |
| Australian born | 1.0 | 1.0 |
| Other English-Speaking Background | 1.71(1.28, 2.29) | 1.63(1.19, 2.23) |
| Europe | 0.85(0.54, 1.32) | 0.86(0.53, 1.38) |
| Asia | 1.22(0.67, 2.24) | 1.34(0.68, 2.65) |
| Other | 1.31(0.53, 3.25) | 1.47(0.55, 3.89) |
| **Education status** |  |  |
| No formal | 1.0 | 1.0 |
| School Cert. | 1.1(0.81, 1.49) | 1.18(0.85, 1.64) |
| Higher school Cert | 1.27(0.85, 1.88) | 1.27(0.83, 1.94) |
| Trade/Apprentice | 0.56(0.31, 1.04) | 0.55(0.29, 1.05) |
| Certificate/Diploma | 1.18(0.84, 1.66) | 1.3(0.9, 1.88) |
| University degree | 1.23(0.82, 1.84) | 1.33(0.86, 2.06) |
| Higher degree | 1.76(1.13, 2.73) | 1.74(1.08, 2.81) |
| **ARIA group** |  |  |
| Major cities | 1.0 | 1.0 |
| Inner regional | 0.77(0.65, 0.93) | 0.78(0.64, 0.94) |
| Outer regional | 0.95(0.74, 1.22) | 0.88(0.67, 1.16) |
| Remote/Very remote | 1.09(0.56, 2.13) | 0.63(0.29, 1.37) |
| **Marital status** |  |  |
| Married | 1.0 | 1.0 |
| Divorced | 2.02(1.57, 2.61) | 1.62(1.23, 2.15) |
| De Facto | 1.62(1.18, 2.22) | 1.39(0.99, 1.95) |
| Widowed | 1.79(1.32, 2.44) | 1.22(0.88, 1.69) |
| Separated | 1.79(1.24, 2.57) | 1.41(0.95, 2.09) |
| Never married | 1.37(0.81, 2.34) | 1.1(0.62, 1.94) |
| **Labour force status** |  |  |
| Employed | 1.0 | 1.0 |
| Unemployed | 1.71(0.97, 2.99) | 1.43(0.76, 2.69) |
| Not in labour force | 1.71(1.49, 1.97) | 1.08(0.92, 1.27) |
| **Regular breast exam** |  |  |
| No | 1.0 | 1.0 |
| Yes | 1.05(0.91, 1.21) | 1.08(0.92, 1.26) |
| **Any chronic disease** |  |  |

|  |  |  |  |
| --- | --- | --- | --- |
|  | No | 1.0 | 1.0 |
|  | Yes | 1.35(1.17, 1.55) | 1.05(0.9, 1.23) |
| **Any cancer** |  |  |  |
|  | No | 1.0 | 1.0 |
|  | Yes | 1.61(1.37, 1.9) | 1.29(1.09, 1.54) |
| **Any psychiatric disorder** |  |  |  |
|  | No | 1.0 | 1.0 |
|  | Yes | 1.34(1.12, 1.59) | 1.19(0.98, 1.44) |
| **Ease of having a mammogram** |  |  |  |
|  | Excellent | 1.0 | 1.0 |
|  | Very good | 1.14(0.98, 1.34) | 1.14(0.96, 1.34) |
|  | Good | 1.41(1.18, 1.69) | 1.53(1.26, 1.85) |
|  | Fair | 2.54(1.92, 3.37) | 3.41(2.51, 4.63) |
|  | Poor | 1.99(1.25, 3.16) | 3.07(1.86, 5.07) |
|  | Don't know | 2.88(1.98, 4.18) | 3.63(2.42, 5.45) |
| **Smoking status** |  |  |  |
|  | never-smoker | 1.0 | 1.0 |
|  | ex-smoker | 1.11(0.92, 1.34) | 1.06(0.87, 1.29) |
|  | smoker | 1.17(0.9, 1.53) | 1.54(1.15, 2.08) |
| **Ever had abnormal pap result** |  |  |  |
|  | No | 1.0 | 1.0 |
|  | Yes | 1.5(1.23, 1.82) | 1.11(0.9, 1.37) |
|  | Don't know | 2.05(1.25, 3.38) | 1.04(0.61, 1.78) |
| **Ever had abnormal mammogram result** |  |  |
| No | 1.0 | 1.0 |
| Yes | 2.57(2.14, 3.08) | 1.71(1.41, 2.08) |
| Don't know | 4.53(1.55, 13.2) | 1.87(0.6, 5.82) |
| **Breast examined by a doctor or nurse** |  |  |
| No | 1.0 | 1.0 |
| Yes | 1.13(0.99, 1.31) | 1.26(1.08, 1.47) |
| **Current hormone replacement therapy** |  |  |
| No | 1.0 | 1.0 |
| Yes | 0.55(0.45, 0.67) | 0.7(0.56, 0.87) |

**Supplementary Table 2.**
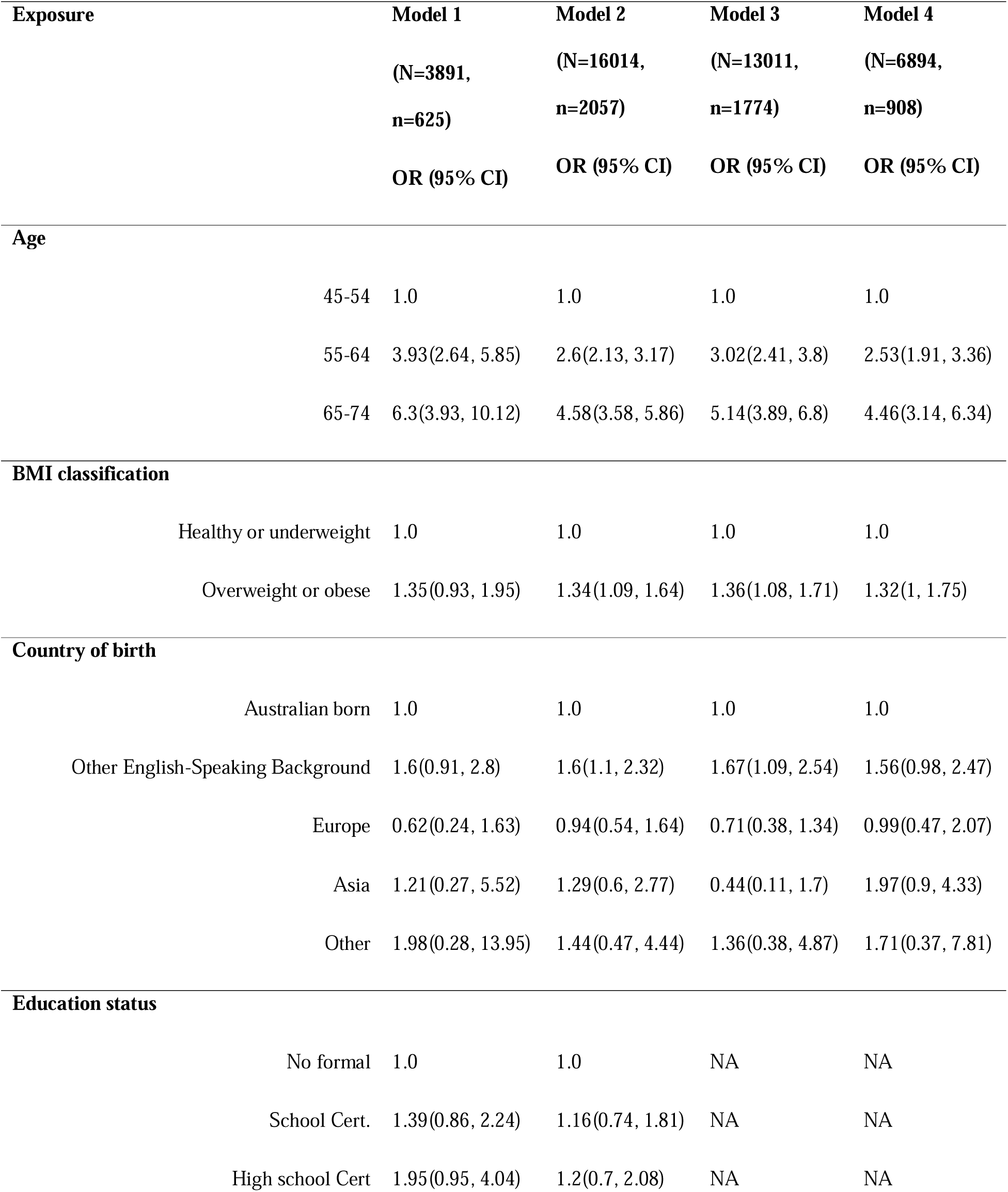
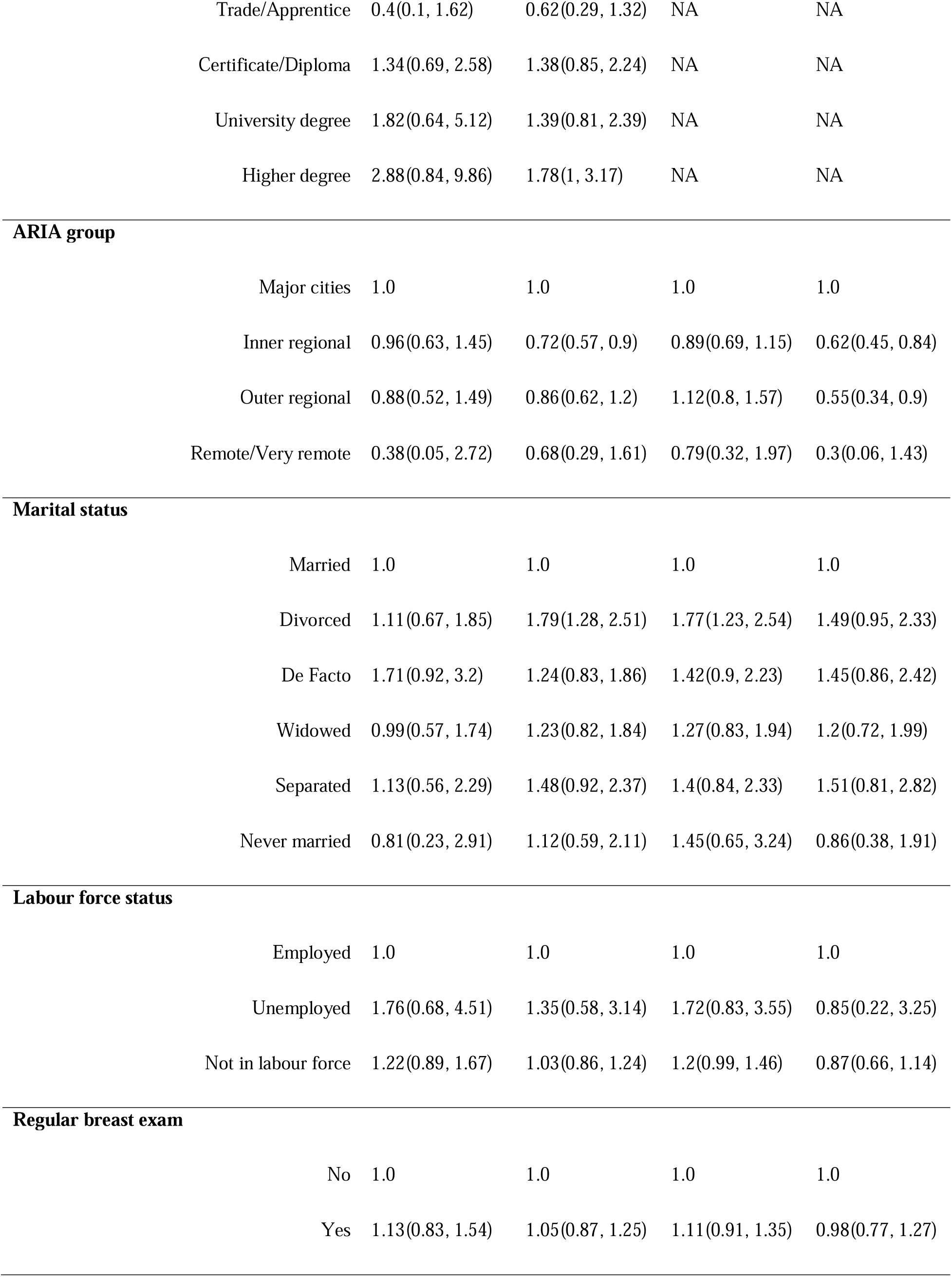
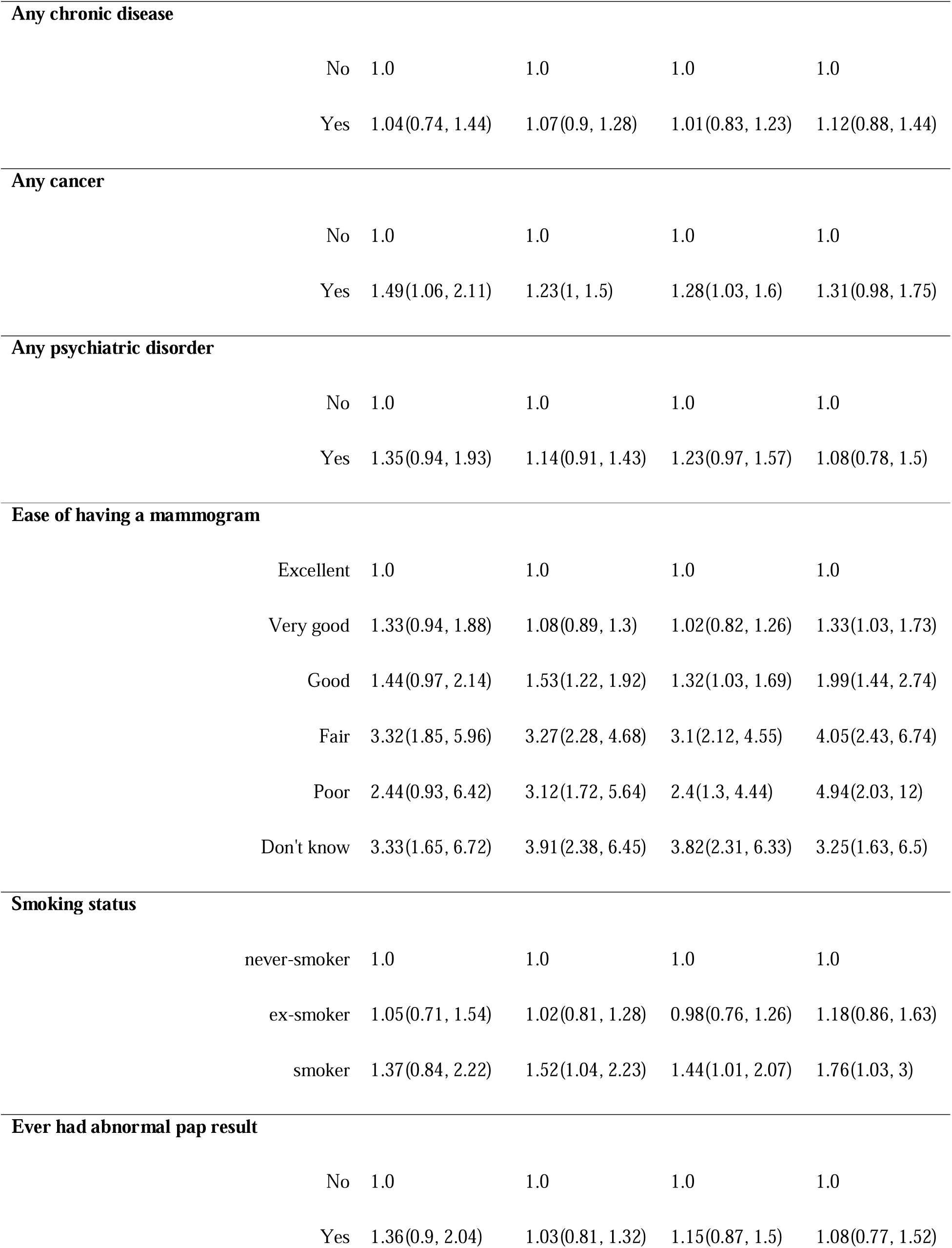
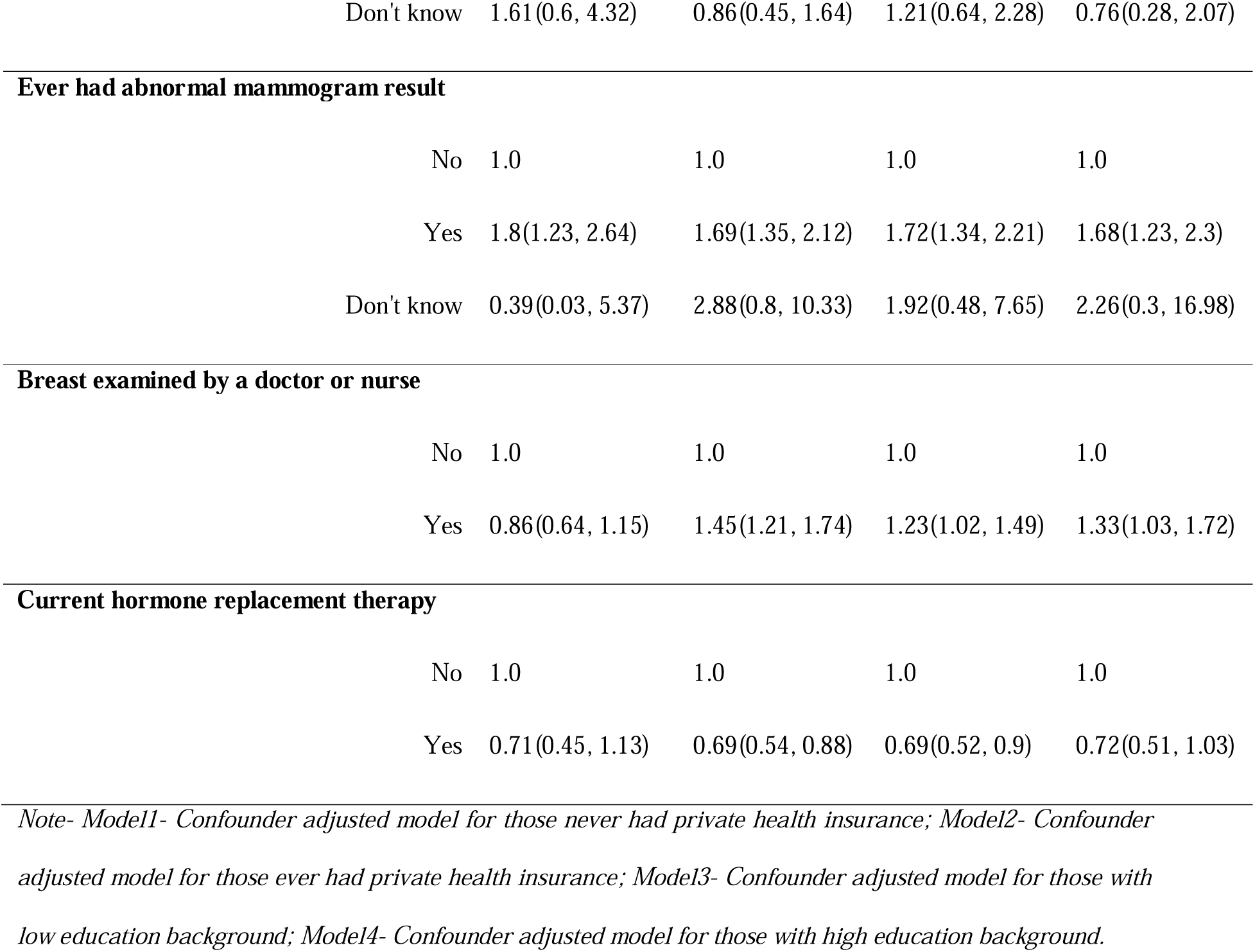
Effect of body mass index category on breast screen attendance, stratified by private health insurance and education status (multilevel logistic regression approach)

| Exposure | Model 1<br>(N=3891,<br>n=625)<br>OR (95% CI) | Model 2<br>(N=16014,<br>n=2057)<br>OR (95% CI) | Model 3<br>(N=13011,<br>n=1774)<br>OR (95% CI) | Model 4<br>(N=6894,<br>n=908)<br>OR (95% CI) |
| --- | --- | --- | --- | --- |
| **Age** |  |  |  |  |
| 45-54 | 1.0 | 1.0 | 1.0 | 1.0 |
| 55-64 | 3.93(2.64, 5.85) | 2.6(2.13, 3.17) | 3.02(2.41, 3.8) | 2.53(1.91, 3.36) |
| 65-74 | 6.3(3.93, 10.12) | 4.58(3.58, 5.86) | 5.14(3.89, 6.8) | 4.46(3.14, 6.34) |
| **BMI classification** |  |  |  |  |
| Healthy or underweight | 1.0 | 1.0 | 1.0 | 1.0 |
| Overweight or obese | 1.35(0.93, 1.95) | 1.34(1.09, 1.64) | 1.36(1.08, 1.71) | 1.32(1, 1.75) |
| **Country of birth** |  |  |  |  |
| Australian born | 1.0 | 1.0 | 1.0 | 1.0 |
| Other English-Speaking Background | 1.6(0.91, 2.8) | 1.6(1.1, 2.32) | 1.67(1.09, 2.54) | 1.56(0.98, 2.47) |
| Europe | 0.62(0.24, 1.63) | 0.94(0.54, 1.64) | 0.71(0.38, 1.34) | 0.99(0.47, 2.07) |
| Asia | 1.21(0.27, 5.52) | 1.29(0.6, 2.77) | 0.44(0.11, 1.7) | 1.97(0.9, 4.33) |
| Other | 1.98(0.28, 13.95) | 1.44(0.47, 4.44) | 1.36(0.38, 4.87) | 1.71(0.37, 7.81) |
| **Education status** |  |  |  |  |
| No formal | 1.0 | 1.0 | NA | NA |
| School Cert. | 1.39(0.86, 2.24) | 1.16(0.74, 1.81) | NA | NA |
| High school Cert | 1.95(0.95, 4.04) | 1.2(0.7, 2.08) | NA | NA |

|  |  |  |  |  |  |
| --- | --- | --- | --- | --- | --- |
|  | Trade/Apprentice | 0.4(0.1, 1.62) | 0.62(0.29, 1.32) | NA | NA |
|  | Certificate/Diploma | 1.34(0.69, 2.58) | 1.38(0.85, 2.24) | NA | NA |
|  | University degree | 1.82(0.64, 5.12) | 1.39(0.81, 2.39) | NA | NA |
|  | Higher degree | 2.88(0.84, 9.86) | 1.78(1, 3.17) | NA | NA |
| **ARIA group** |  |  |  |  |  |
|  | Major cities | 1.0 | 1.0 | 1.0 | 1.0 |
|  | Inner regional | 0.96(0.63, 1.45) | 0.72(0.57, 0.9) | 0.89(0.69, 1.15) | 0.62(0.45, 0.84) |
|  | Outer regional | 0.88(0.52, 1.49) | 0.86(0.62, 1.2) | 1.12(0.8, 1.57) | 0.55(0.34, 0.9) |
|  | Remote/Very remote | 0.38(0.05, 2.72) | 0.68(0.29, 1.61) | 0.79(0.32, 1.97) | 0.3(0.06, 1.43) |
| **Marital status** |  |  |  |  |  |
|  | Married | 1.0 | 1.0 | 1.0 | 1.0 |
|  | Divorced | 1.11(0.67, 1.85) | 1.79(1.28, 2.51) | 1.77(1.23, 2.54) | 1.49(0.95, 2.33) |
|  | De Facto | 1.71(0.92, 3.2) | 1.24(0.83, 1.86) | 1.42(0.9, 2.23) | 1.45(0.86, 2.42) |
|  | Widowed | 0.99(0.57, 1.74) | 1.23(0.82, 1.84) | 1.27(0.83, 1.94) | 1.2(0.72, 1.99) |
|  | Separated | 1.13(0.56, 2.29) | 1.48(0.92, 2.37) | 1.4(0.84, 2.33) | 1.51(0.81, 2.82) |
|  | Never married | 0.81(0.23, 2.91) | 1.12(0.59, 2.11) | 1.45(0.65, 3.24) | 0.86(0.38, 1.91) |
| **Labour force status** |  |  |  |  |  |
|  | Employed | 1.0 | 1.0 | 1.0 | 1.0 |
|  | Unemployed | 1.76(0.68, 4.51) | 1.35(0.58, 3.14) | 1.72(0.83, 3.55) | 0.85(0.22, 3.25) |
|  | Not in labour force | 1.22(0.89, 1.67) | 1.03(0.86, 1.24) | 1.2(0.99, 1.46) | 0.87(0.66, 1.14) |
| **Regular breast exam** |  |  |  |  |  |
|  | No | 1.0 | 1.0 | 1.0 | 1.0 |
|  | Yes | 1.13(0.83, 1.54) | 1.05(0.87, 1.25) | 1.11(0.91, 1.35) | 0.98(0.77, 1.27) |
| **Any chronic disease** |  |  |  |  |
| No | 1.0 | 1.0 | 1.0 | 1.0 |
| Yes | 1.04(0.74, 1.44) | 1.07(0.9, 1.28) | 1.01(0.83, 1.23) | 1.12(0.88, 1.44) |
| **Any cancer** |  |  |  |  |
| No | 1.0 | 1.0 | 1.0 | 1.0 |
| Yes | 1.49(1.06, 2.11) | 1.23(1, 1.5) | 1.28(1.03, 1.6) | 1.31(0.98, 1.75) |
| **Any psychiatric disorder** |  |  |  |  |
| No | 1.0 | 1.0 | 1.0 | 1.0 |
| Yes | 1.35(0.94, 1.93) | 1.14(0.91, 1.43) | 1.23(0.97, 1.57) | 1.08(0.78, 1.5) |
| **Ease of having a mammogram** |  |  |  |  |
| Excellent | 1.0 | 1.0 | 1.0 | 1.0 |
| Very good | 1.33(0.94, 1.88) | 1.08(0.89, 1.3) | 1.02(0.82, 1.26) | 1.33(1.03, 1.73) |
| Good | 1.44(0.97, 2.14) | 1.53(1.22, 1.92) | 1.32(1.03, 1.69) | 1.99(1.44, 2.74) |
| Fair | 3.32(1.85, 5.96) | 3.27(2.28, 4.68) | 3.1(2.12, 4.55) | 4.05(2.43, 6.74) |
| Poor | 2.44(0.93, 6.42) | 3.12(1.72, 5.64) | 2.4(1.3, 4.44) | 4.94(2.03, 12) |
| Don't know | 3.33(1.65, 6.72) | 3.91(2.38, 6.45) | 3.82(2.31, 6.33) | 3.25(1.63, 6.5) |
| **Smoking status** |  |  |  |  |
| never-smoker | 1.0 | 1.0 | 1.0 | 1.0 |
| ex-smoker | 1.05(0.71, 1.54) | 1.02(0.81, 1.28) | 0.98(0.76, 1.26) | 1.18(0.86, 1.63) |
| smoker | 1.37(0.84, 2.22) | 1.52(1.04, 2.23) | 1.44(1.01, 2.07) | 1.76(1.03, 3) |
| **Ever had abnormal pap result** |  |  |  |  |
| No | 1.0 | 1.0 | 1.0 | 1.0 |
| Yes | 1.36(0.9, 2.04) | 1.03(0.81, 1.32) | 1.15(0.87, 1.5) | 1.08(0.77, 1.52) |

|  |  |  |  |  |  |
| --- | --- | --- | --- | --- | --- |
|  | Don't know | 1.61(0.6, 4.32) | 0.86(0.45, 1.64) | 1.21(0.64, 2.28) | 0.76(0.28, 2.07) |
| **Ever had abnormal mammogram result** |  |  |  |  |  |
|  | No | 1.0 | 1.0 | 1.0 | 1.0 |
|  | Yes | 1.8(1.23, 2.64) | 1.69(1.35, 2.12) | 1.72(1.34, 2.21) | 1.68(1.23, 2.3) |
|  | Don't know | 0.39(0.03, 5.37) | 2.88(0.8, 10.33) | 1.92(0.48, 7.65) | 2.26(0.3, 16.98) |
| **Breast examined by a doctor or nurse** |  |  |  |  |  |
|  | No | 1.0 | 1.0 | 1.0 | 1.0 |
|  | Yes | 0.86(0.64, 1.15) | 1.45(1.21, 1.74) | 1.23(1.02, 1.49) | 1.33(1.03, 1.72) |
| **Current hormone replacement therapy** |  |  |  |  |  |
|  | No | 1.0 | 1.0 | 1.0 | 1.0 |
|  | Yes | 0.71(0.45, 1.13) | 0.69(0.54, 0.88) | 0.69(0.52, 0.9) | 0.72(0.51, 1.03) |

*Note- Model1- Confounder adjusted model for those never had private health insurance; Model2- Confounder adjusted model for those ever had private health insurance; Model3- Confounder adjusted model for those with low education background; Model4- Confounder adjusted model for those with high education background.*

